# Trends in Influenza-Associated Lower Respiratory Tract Infection Burden and Influenza Vaccination Coverage among Adults Aged ≥55 Years in China, 1990–2023

**DOI:** 10.64898/2026.07.11.26357807

**Authors:** Min Liu, Ruochan Chen

**Author notes:** Corresponding author: Ruochan Chen.

## Abstract

**Background:** Influenza-associated lower respiratory tract infections (LRTI) impose substantial disease burden on older populations globally. While China has experienced rapid aging, comprehensive assessment of long-term trends in influenza-associated LRTI burden and vaccination coverage among adults aged ≥55 years remains limited.

**Methods:** We extracted age-standardized mortality and disability-adjusted life years (DALYs) rates of influenza-associated LRTI among Chinese adults aged ≥55 years from the Global Burden of Disease Study 2023 (GBD 2023). Estimated annual percentage change (EAPC) and age-period-cohort (APC) models were employed to analyze long-term trends (1990–2023). Using the China Health and Retirement Longitudinal Study 2020 (CHARLS 2020, n=13,815), we performed multivariable logistic regression to identify factors associated with influenza vaccination uptake. Spearman correlation assessed ecological associations between GBD age-specific mortality rates and CHARLS vaccination coverage across nine age groups.

**Results:** Age-standardized mortality rates declined from 4.09 to 0.31 per 100,000 (EAPC=-8.02%, 95%CI: −9.08% to −6.95%), and DALYs rates from 177.63 to 6.06 per 100,000 (EAPC=-10.67%), representing >96% reductions from 1990 to 2023. Deaths dropped sharply by 59.6% in 2020, consistent with protective effects of non-pharmaceutical interventions during COVID-19. The ≥85-year age group showed the slowest improvement (EAPC=-5.63%). Influenza vaccination coverage in CHARLS 2020 was only 15.1%. Older age (≥85 years: OR=0.65, 95%CI: 0.46–0.92) and higher educational attainment (college or above: OR=0.34, 95%CI: 0.20–0.56) were independently associated with lower vaccination rates, though the small sample size in the highest education group (n=68) warrants cautious interpretation. Ecological analysis revealed a significant negative correlation between GBD mortality rates and CHARLS vaccination coverage (r=-0.700, p=0.036, n=9 age groups), indicating a “protection mismatch” phenomenon. Raising vaccination coverage to 75% (WHO target) could prevent approximately 2,041 deaths annually under moderate vaccine efficacy assumptions, with ≥75-year-olds contributing 82.7% of preventable deaths.

**Conclusions:** Influenza-associated LRTI burden has declined substantially among Chinese adults aged ≥55 years, but improvement is slowest in the oldest age groups. Vaccination coverage remains critically insufficient with a persistent “protection mismatch” where those at highest mortality risk have lowest coverage. Adults aged ≥75 years face a “triple vulnerability” of high mortality, low vaccination coverage, and high COPD comorbidity prevalence, and should be prioritized for vaccination to maximize preventable mortality benefits.

## BACKGROUND

Each year, influenza accounts for an estimated 1 billion infections globally, 3–5 million of which progress to severe illness; respiratory deaths attributable to the virus range from 290,000 to 650,000 [1,2]. The clinical burden extends beyond direct viral injury — influenza disrupts airway epithelial integrity and blunts local immune defenses, creating conditions that favor secondary bacterial pneumonia and, in turn, driving the majority of influenza-related fatalities. Three organisms dominate these co-infections: Streptococcus pneumoniae, Staphylococcus aureus, and Haemophilus influenzae. When influenza acts in concert with any of these pathogens, case fatality rates rise well above what either agent causes alone — a synergistic effect that is particularly consequential in patients with underlying lung disease [3,4].

Among all age groups, older adults bear a disproportionate burden of influenza-associated LRTI. Immunosenescence and inflammaging — the chronic low-grade inflammatory state that accompanies aging — erode the capacity to clear virus efficiently, leaving older individuals far more susceptible to pneumonia, hospitalization, and death than their younger counterparts [5,6]. The clinical gap is not subtle: compared with younger adults, older patients infected with influenza are substantially more likely to progress to pneumonia requiring mechanical ventilation and to die within 30 days. Chronic comorbidities compound this vulnerability. COPD, cardiovascular disease, diabetes mellitus, and chronic kidney disease each independently impair physiological reserve; together, they create the permissive microenvironments that facilitate secondary bacterial invasion. Data from the Global Burden of Disease (GBD) study confirm that adults aged ≥65 years account for the vast majority of influenza-associated LRTI deaths worldwide [7].

China, the world’s most populous nation, is experiencing rapid population aging. In 2020, 264 million Chinese were aged ≥60 years (18.7% of the population), projected to exceed 400 million by 2050. The “4-2-1” family structure—in which one child supports two parents and four grandparents—constrains informal caregiving capacity, while rapid urbanization disrupts traditional multigenerational support networks that historically buffered health risks in older adults [8]. This accelerating demographic transition intensifies concerns about influenza-associated LRTI burden. While previous studies have documented declining LRTI burden in China during 1990–2019, national rates remain above global averages [9]. Adults aged ≥55 years represent a critical life stage characterized by accelerating functional decline and chronic disease onset, coinciding with labor force retirement—an age threshold adopted by major international aging cohort studies. However, comprehensive long-term trend analyses of influenza-associated LRTI burden specifically targeting this population using the latest GBD 2023 data remain scarce [10].

Vaccination remains the cornerstone of influenza prevention [11]. In older adults, it cuts hospitalization risk by 40–60% and reduces mortality by roughly half — gains that are clinically meaningful given the baseline risk this population carries. High-dose and adjuvanted formulations, developed specifically to overcome the blunted immune response of senescent immune systems, outperform standard-dose vaccines on both immunogenicity and real-world effectiveness [12]. Yet in China, uptake has remained stubbornly low [13]. Coverage in adults aged ≥60 years has historically fluctuated between 2% and 20%, well short of the WHO’s 75% target and the national programme’s more modest goal of ≥60% — with wide interprovincial gaps driven by uneven subsidy policies, unequal access to primary care, and low public awareness. Identifying who gets vaccinated and, critically, who does not — and why — is a prerequisite for designing interventions that actually reach the people most at risk [14].

The China Health and Retirement Longitudinal Study (CHARLS) provides one of the richest individual-level datasets on aging in China, covering health status, health behaviors, and socioeconomic circumstances across a nationally representative sample [15]. Its 2020 wave — fielded between July and December 2020, deep into the first year of the COVID-19 pandemic — included a dedicated supplementary module on influenza vaccination, respiratory illness, and healthcare-seeking. The timing was fortuitous in a research sense: the co-occurrence of mass NPI deployment and detailed vaccination records creates an unusual window for examining both how NPIs shaped influenza transmission and what drove vaccine uptake under pandemic conditions.

This study integrates GBD 2023 macro-level data with CHARLS 2020 micro-level survey data to: (1) describe long-term trends (1990–2023) in influenza-associated LRTI mortality and DALYs rates among Chinese adults aged ≥55 years, decomposing age, period, and cohort effects through APC modelling; (2) analyse baseline characteristics and determinants of influenza vaccination using CHARLS 2020; (3) conduct GBD–CHARLS joint ecological analysis to elucidate associations between macro-level mortality burden and micro-level population characteristics; and (4) quantify potential preventable deaths achievable by raising vaccination coverage to national targets, providing evidence to inform targeted prevention strategies.

## METHODS

### Study Design and Data Sources

This study employed a dual-data-source approach combining macro-level disease burden estimates with micro-level population survey data.

GBD 2023: We extracted mortality counts, mortality rates, and disability-adjusted life years (DALYs) rates for influenza-associated LRTI among Chinese adults aged ≥55 years from 1990 to 2023, stratified by sex and nine 5-year age groups. Data were obtained from the publicly available GBD 2023 database (https://vizhub.healthdata.org/gbd-results).

CHARLS 2020: CHARLS is a nationally representative longitudinal cohort study covering 28 Chinese provinces. For this analysis, we included CHARLS 2020 respondents aged ≥55 years with complete data on key variables. After excluding individuals with missing age information, 13,815 participants (age range: 55–108 years) were included.

### Statistical Analysis

Long-term trend analysis: Estimated annual percentage change (EAPC) was calculated to describe long-term trends in age-standardized mortality and DALYs rates from 1990 to 2023 [15,16]. Age-period-cohort (APC) models using the intrinsic estimator method were employed to decompose age, period, and cohort effects [17,18].

Baseline characteristics: Continuous variables were presented as mean±SD; categorical variables as frequencies and proportions. Chi-square tests compared group differences (p<0.05).

Multivariable logistic regression: We constructed multivariable logistic regression models with influenza vaccination as the dependent variable, and age group (reference: 55–64 years), sex (reference: male), residence (reference: urban), smoking history, COPD, and educational attainment (reference: illiterate/primary school) as independent variables. Odds ratios (OR) and 95% confidence intervals (CI) were calculated. As this was an exploratory analysis, no multiple comparison corrections were applied; results should be interpreted cautiously.

Ecological correlation analysis: Spearman rank correlation assessed associations between GBD age-specific mortality rates and CHARLS age-group vaccination coverage (n=9 age groups). Note that ecological study designs cannot infer individual-level causal relationships [19].

Preventable death estimation: Assuming vaccination coverage increased to the WHO-recommended target of 75% for older adults [20], we estimated preventable deaths using: Preventable deaths = Deaths × max(75% − current coverage, 0) × VE. Based on meta-analyses [21,22], we modeled three vaccine efficacy scenarios: conservative (VE=40%), moderate (VE=50%), and optimistic (VE=60%).

All statistical analyses were performed using Python 3.12 and R 4.5. Two-sided p<0.05 was considered statistically significant.

### Ethics

CHARLS was approved by the Biomedical Ethics Committee of Peking University (IRB00001052-11015); all participants provided informed consent. This study used publicly available de-identified data requiring no additional ethical review. GBD 2023 data are publicly available with no ethical restrictions.

## RESULTS

### Disease Burden Trends, 1990–2023

From 1990 to 2023, age-standardized mortality rates for influenza-associated LRTI among Chinese adults aged ≥55 years declined from 4.09 per 100,000 (95% UI: 3.31–4.94) to 0.31 per 100,000 (95% UI: 0.15–0.54), with EAPC=-8.02% (95%CI: −9.08% to −6.95%). DALYs rates declined from 177.63 per 100,000 to 6.06 per 100,000, with EAPC=-10.67% (95%CI: −11.77% to −9.55%), representing >96% reductions for both metrics (Table 1).

**Table 1.**
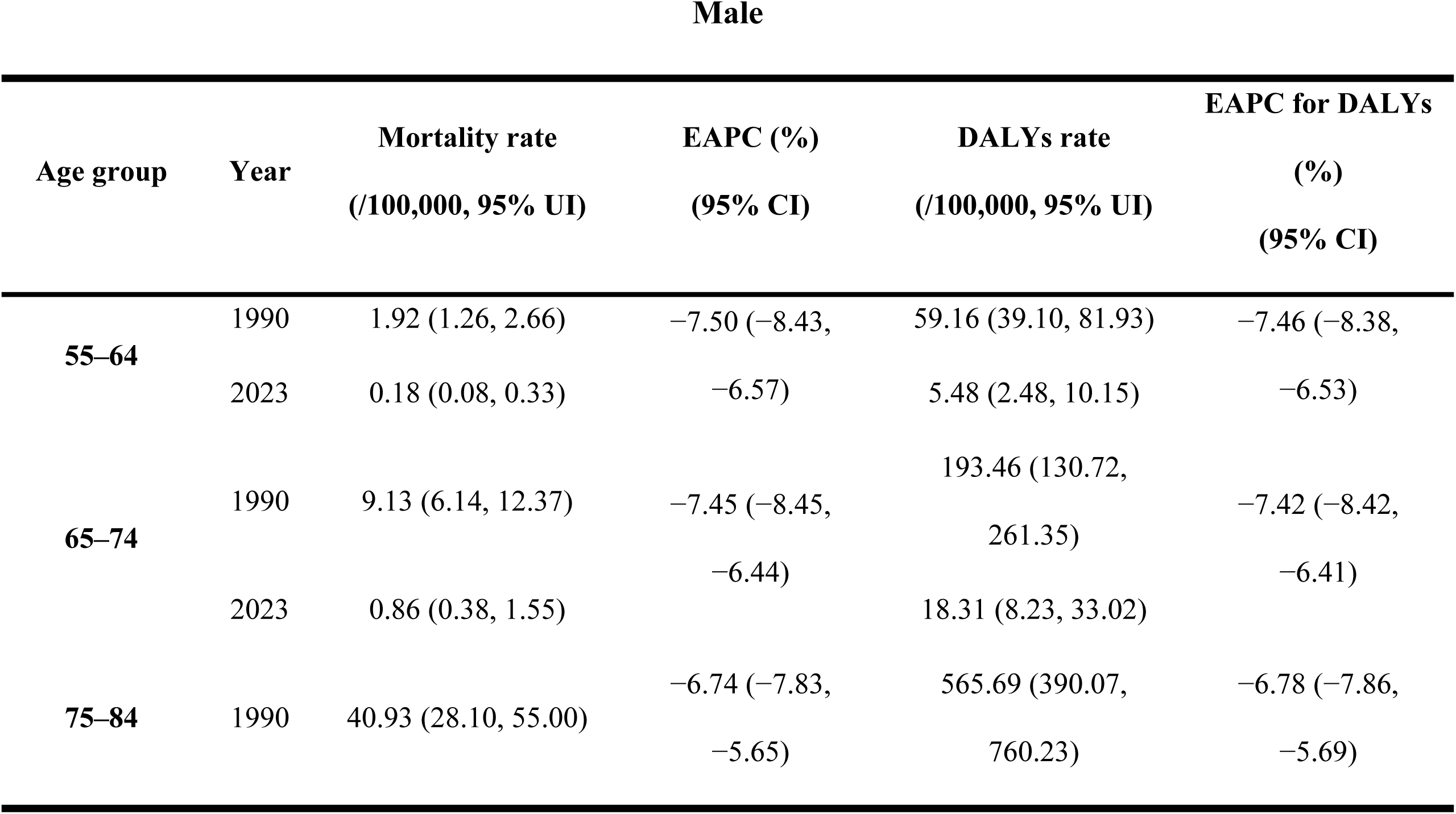
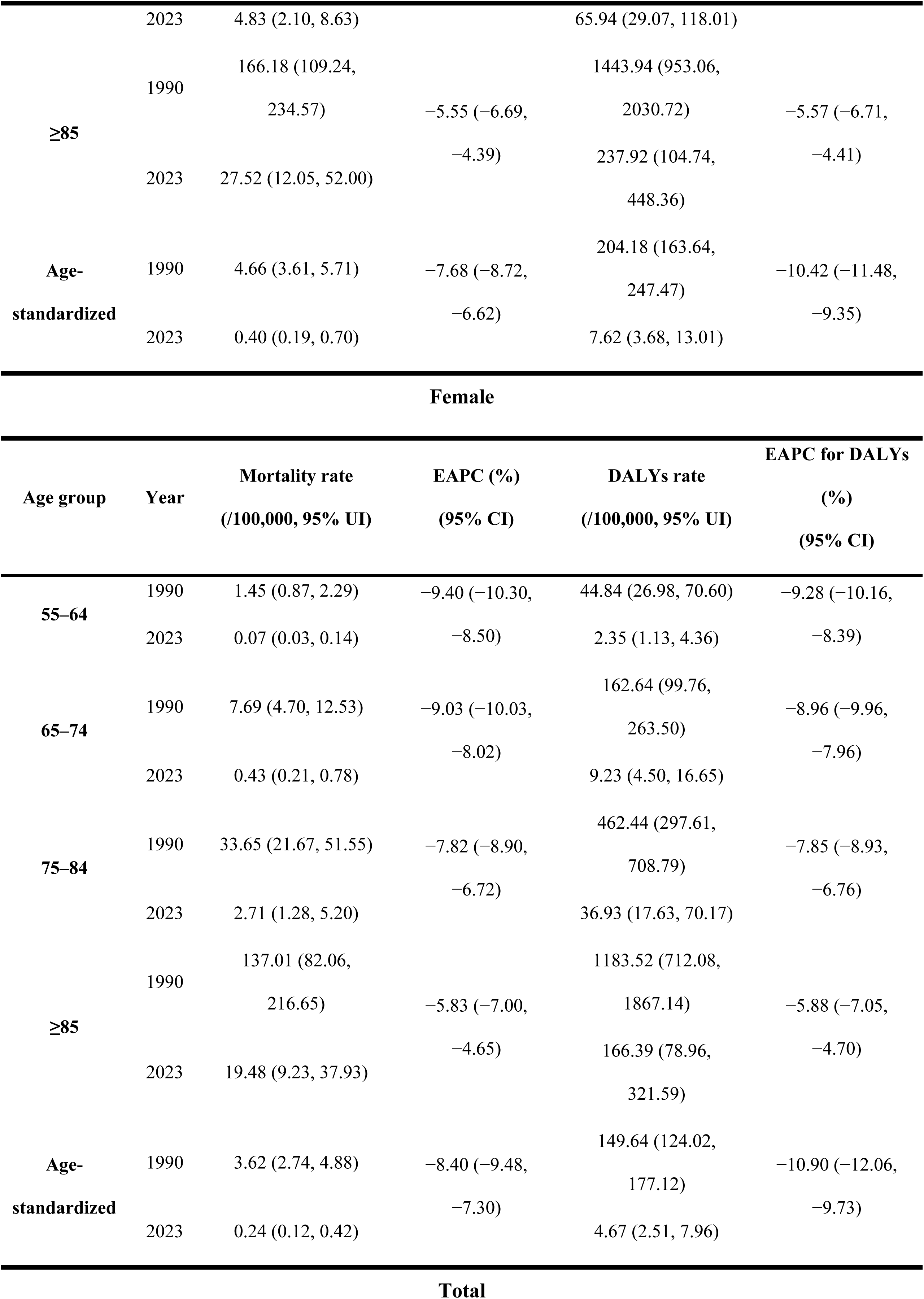
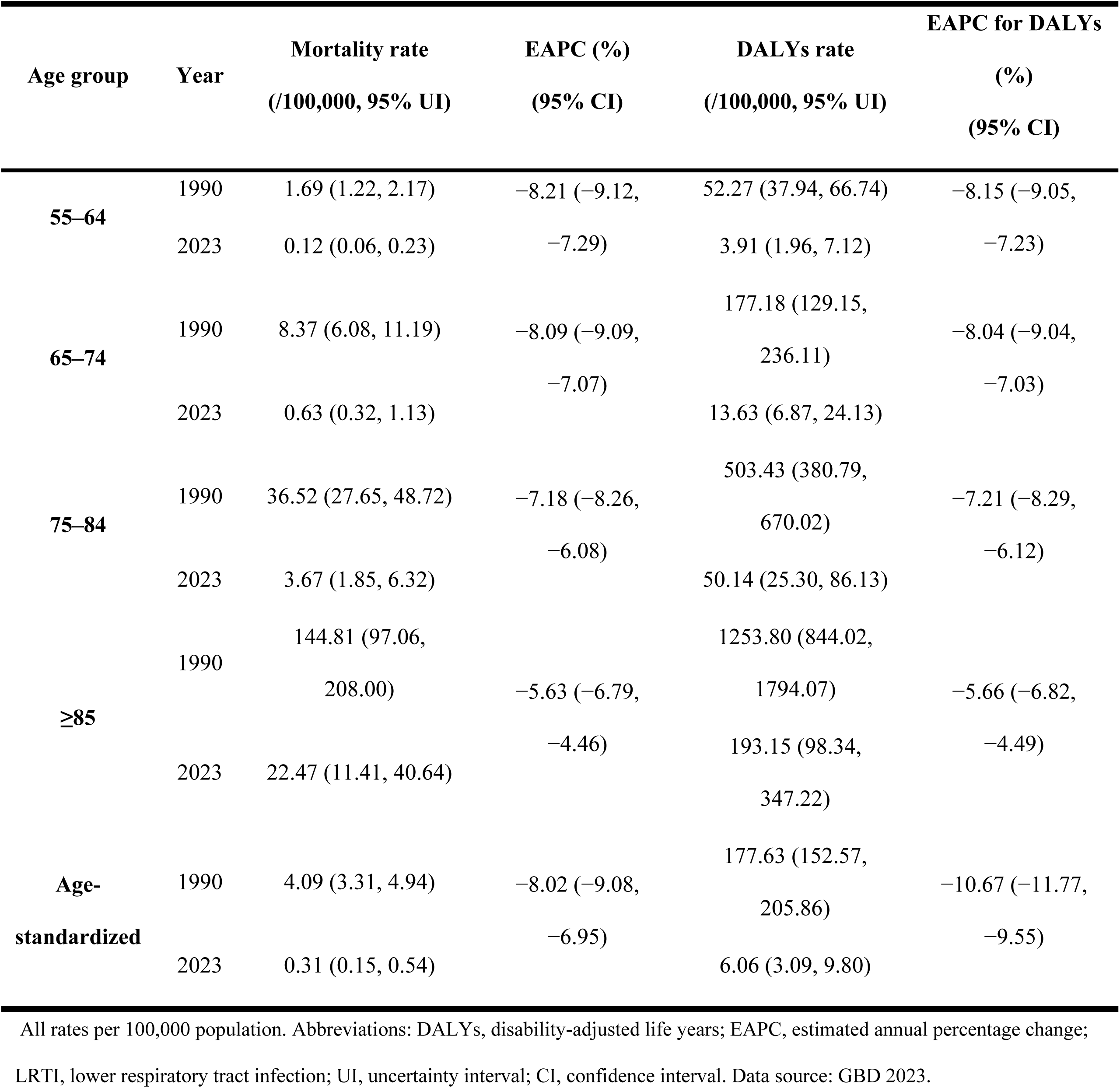
Mortality rates and DALYs rates of influenza-associated LRTI among adults aged ≥55 years in China, by age group and sex, 1990 and 2023.

Males had consistently higher age-standardized mortality rates than females across all years, but females showed slightly greater percentage decline (EAPC: −8.40% vs −7.68%). Mortality rates increased significantly with age across all age groups. The ≥85-year group showed the slowest improvement (EAPC=-5.63%, 95%CI: −6.79% to −4.46%), while the 55–64-year group declined most rapidly (EAPC=-8.21%).

Turning to sex-specific patterns (Figure 1), female deaths dropped from 6,631 in 1990 to 2,543 in 2023; for males, the corresponding figures were 5,778 and 3,265. One shift worth pointing out is that male deaths began to exceed female deaths after 2019—a reversal from earlier years. When we look at age-standardized mortality rates, females experienced a slightly steeper decline than males (93.4% vs 91.4%), although male rates remained higher throughout the entire study period. Both death counts and standardized rates reached their lowest point in 2020, then edged up modestly in 2023.

**Figure 1.**
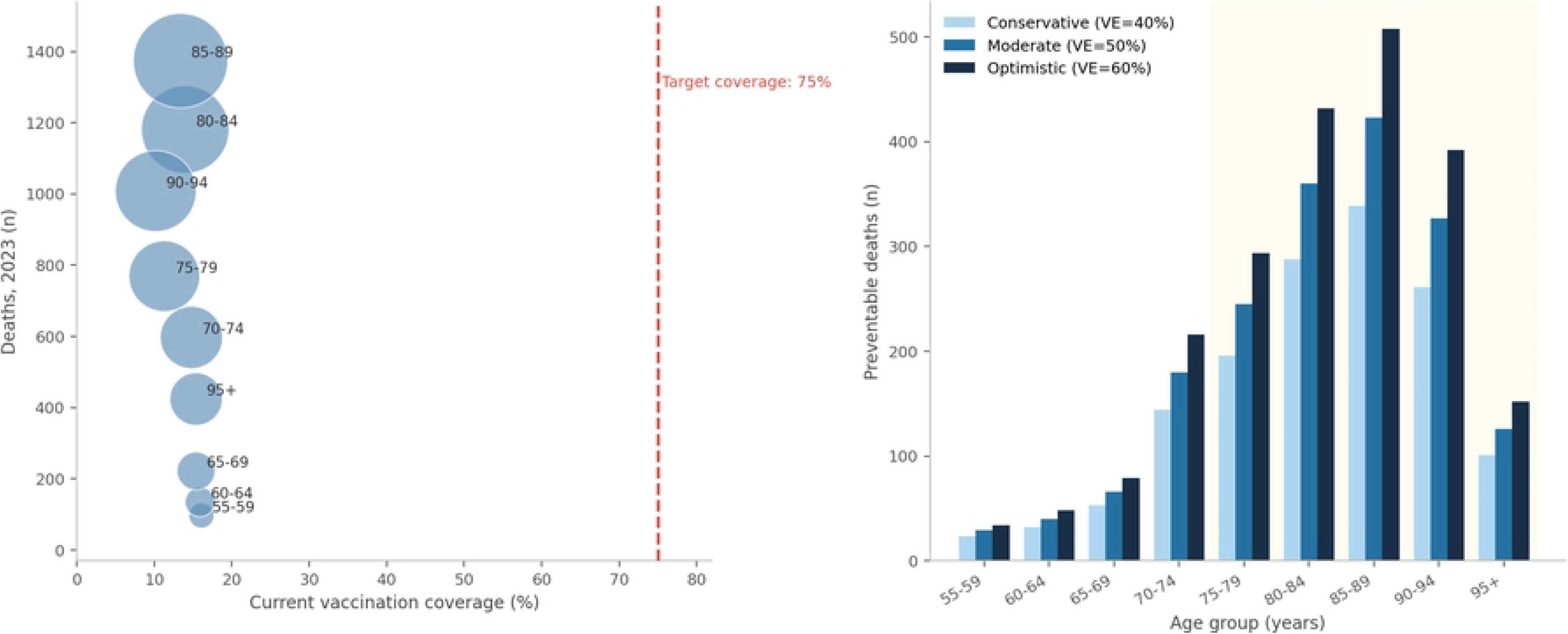
Temporal trends in influenza-associated LRTI burden among adults aged ≥55 years in China, 1990–2023. A: Mortality trends. B: DALYs trends. Bars, absolute counts; lines, age-standardized rates with 95% UI.

### Age-Period-Cohort Model Analysis

Several lines of evidence from the APC model point to a sustained mortality decline (Figure 2). We observed a negative net drift—consistent with the overall downward trend in mortality. The longitudinal age curves rose monotonically with age and, notably, the slope steepened after age 75. What caught our attention was that actual mortality in the ≥85-year group overshot the linear model’s predictions, which we interpret as evidence of additional risk accumulation among the oldest-old.

**Figure 2.**
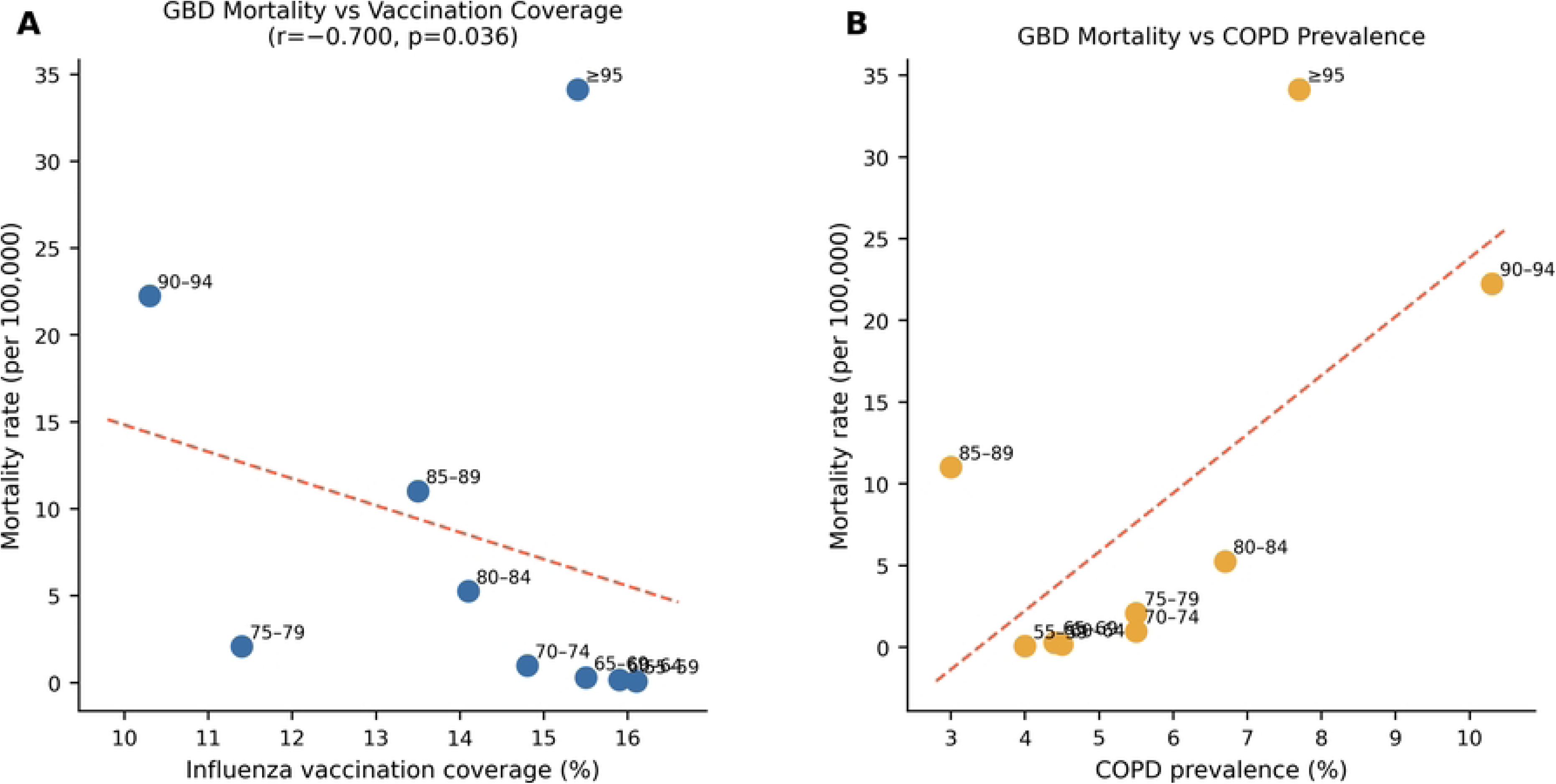
Age-period-cohort (APC) model decomposition of influenza-associated LRTI mortality trends. A: Period effects; B: Cohort effects; C: Local drifts by age group. Reference period: 2007.5; reference cohort: 1930. Error bars, 95% CI

Period effects deviated sharply downward around 2020—coinciding with the 59.6% drop in deaths we described earlier—and this pattern strongly suggests a protective role for non-pharmaceutical interventions during the COVID-19 pandemic. Local drifts were negative across every age group, though the declines were most pronounced in the 55–74-year range and least so among those aged ≥75 years.

### CHARLS 2020 Baseline Characteristics

Our analytical sample comprised 13,815 participants aged 55 years or older, with a mean age of 66.1 years (SD 8.0) (Table 2). The age distribution skewed toward the younger end of the spectrum—nearly half (47.1%) were in the 55–64 bracket, while only 15.8% had reached 75 years or beyond. Sex balance was nearly even, with women accounting for 51.8%. The overwhelming majority (89.2%) lived in urban settings, and educational levels were generally low: 80.0% had no more than a junior high school education. Smoking history was reported by 41.6% of the sample.

**Table 2.** Baseline characteristics of CHARLS 2020 participants aged ≥55 years (N=13,815)

| Characteristic | n | % |
| --- | --- | --- |
| **Age (years), mean±SD** | 66.1±8.0 | — |
| **Age group** |  |  |
| 55–64 years | 6,512 | 47.1 |
| 65–74 years | 5,113 | 37.0 |
| 75–84 years | 1,852 | 13.4 |
| ≥85 years | 338 | 2.4 |
| **Sex** |  |  |
| Male | 6,653 | 48.2 |
| Female | 7,162 | 51.8 |
| **Residence** |  |  |
| Urban | 6,898 | 49.9 |
| Rural | 6,917 | 50.1 |
| **Education** |  |  |
| Illiterate/semi-literate | 3,471 | 25.1 |
| Primary | 2,682 | 19.4 |
| Middle school | 4,908 | 35.5 |
| High school | 1,422 | 10.3 |
| College and above | 68 | 0.5 |
| Missing | 1,283 | 9.3 |
| **Smoking history** | 5,753 | 41.6 |
| **COPD** | 648 | 4.7 |
| **Influenza vaccination (past year)** | 2,090 | 15.1 |

Turning to the key health-related measures, the prevalence of COPD stood at 4.7% overall, whereas the self-reported influenza vaccination rate in the preceding year was notably low—just 15.1% (COPD: physician-diagnosed (CHARLS da003_5_=1). Vaccination: past year (va007=1).

Table 3). When we stratified by age, a clear gradient emerged: COPD prevalence climbed with advancing age, peaking at 5.9% among those aged 75–84 years. Vaccination coverage, by contrast, moved in the opposite direction—16.0% in the youngest age group (55–64) versus 12.3% in the oldest (75–84), a difference that was statistically significant (p=0.001). Perhaps the most striking pattern, however, involved education: coverage was highest among illiterate or semi-literate participants (18.1%) and lowest among those with a college education (4.4%, p<0.001)—a gradient that runs counter to what one might typically expect. The age- and education-related patterns described above are visually confirmed in Figure 3, which plots the same data to highlight these contrasting gradients.

**Figure 3.**
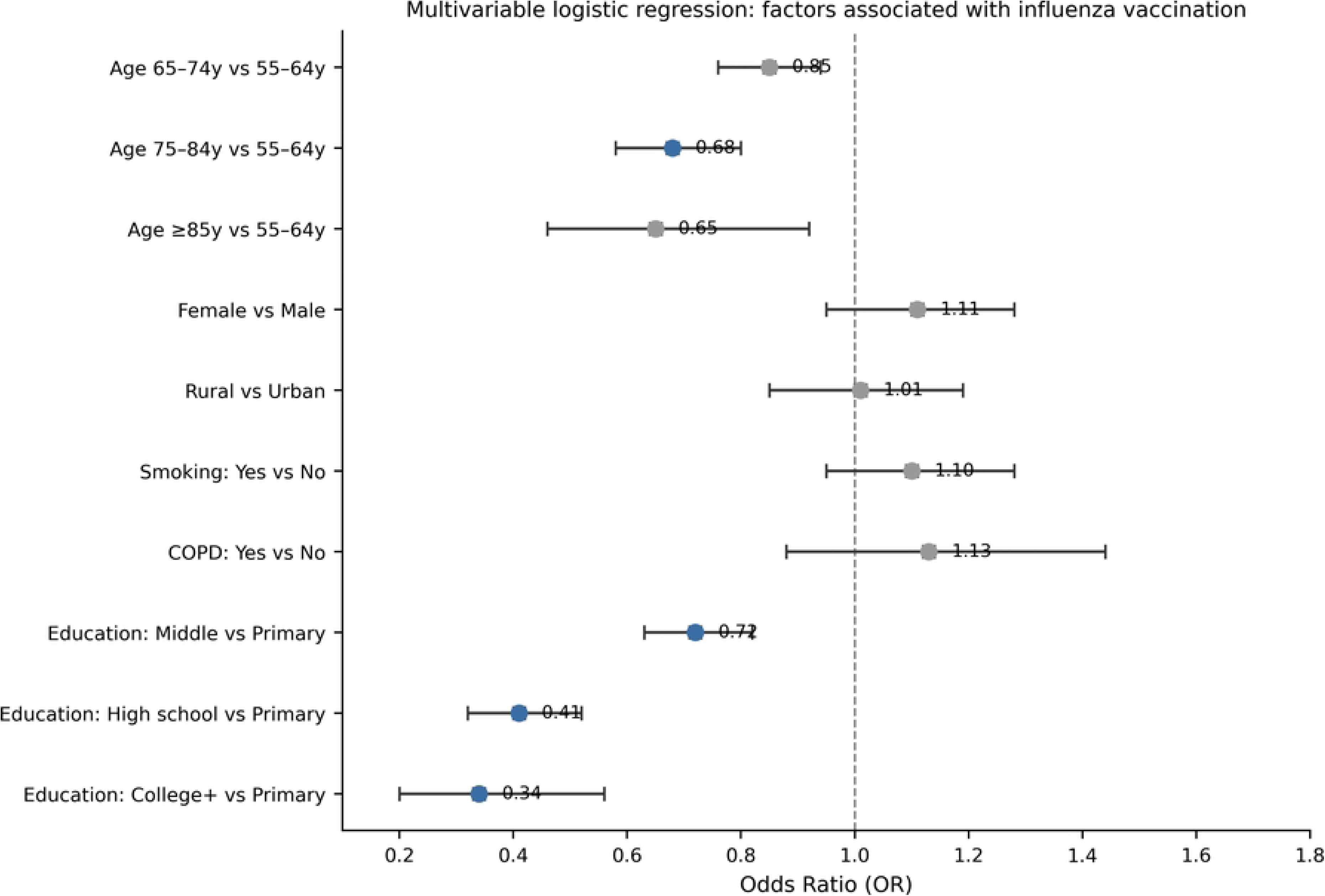
Age-stratified influenza vaccination coverage and COPD prevalence among CHARLS 2020 participants (N=13,815). P=0.001 across age group

**Table 3.** Key characteristics of CHARLS 2020 participants by age group.

| Age group | n | Female (%) | Rural (%) | Smoking (%) | COPD | Vaccination |
| --- | --- | --- | --- | --- | --- | --- |

|  |  |  |  |  | (%) | (%) |
| --- | --- | --- | --- | --- | --- | --- |
| 55–64y | 6,512 | 51.6 | 48.8 | 40.9 | 4.3 | 16.0 |
| 65–74y | 5,113 | 51.4 | 51.4 | 42.4 | 4.8 | 15.2 |
| 75–84y | 1,852 | 52.6 | 50.7 | 43.5 | 5.9 | 12.3 |
| ≥85y | 338 | 59.2 | 52.1 | 34.6 | 4.4 | 13.0 |
| **Total** | **13,815** | **51.8** | **50.1** | **41.6** | **4.7** | **15.1** |

P=0.001 for vaccination coverage across age groups (chi-square test).

### Determinants of Influenza Vaccination

When we ran a multivariable logistic regression to examine factors independently associated with vaccination status, age emerged as a strong predictor. Relative to the 55–64-year group, the odds of being vaccinated dropped progressively with advancing age—ORs were 0.85 (95% CI: 0.76–0.94) for those aged 65–74, 0.68 (95% CI: 0.58–0.80) for 75–84-year-olds, and 0.65 (95% CI: 0.46–0.92) for the oldest group (≥85 years), with all comparisons reaching statistical significance (all p < 0.05).

The education–vaccination relationship, however, followed a rather unexpected pattern. Higher educational attainment was paradoxically associated with lower vaccination odds. Compared with participants who had no more than a primary school education, the adjusted ORs were 0.72 for junior high school graduates, 0.41 for senior high school, and 0.34 (95% CI: 0.20–0.56) for those with a college degree or above (p < 0.001 across all levels). That said, we would caution against over-interpreting the college group estimate—it is based on only 68 individuals, so the precision is inherently limited. Sex, urban/rural residence, and smoking history, on the other hand, showed no statistically meaningful associations with vaccination. The full regression output is provided in Table 4, and the corresponding ORs with 95% CIs are visualized in Figure 4.

**Figure 4.**
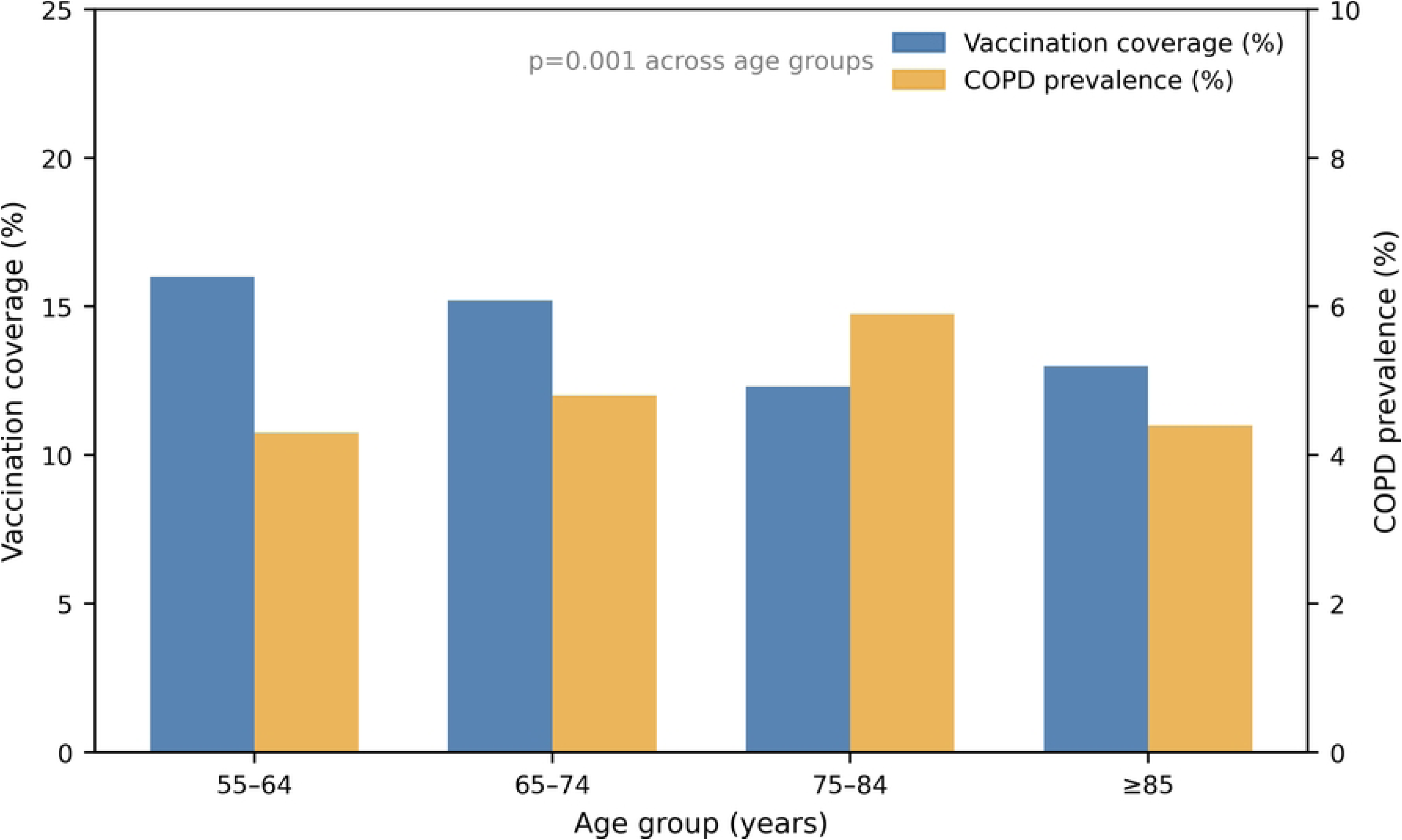
Forest plot of multivariable logistic regression for factors associated with influenza vaccination (CHARLS 2020). Blue = significant (p<0.05).

**Table 4.**
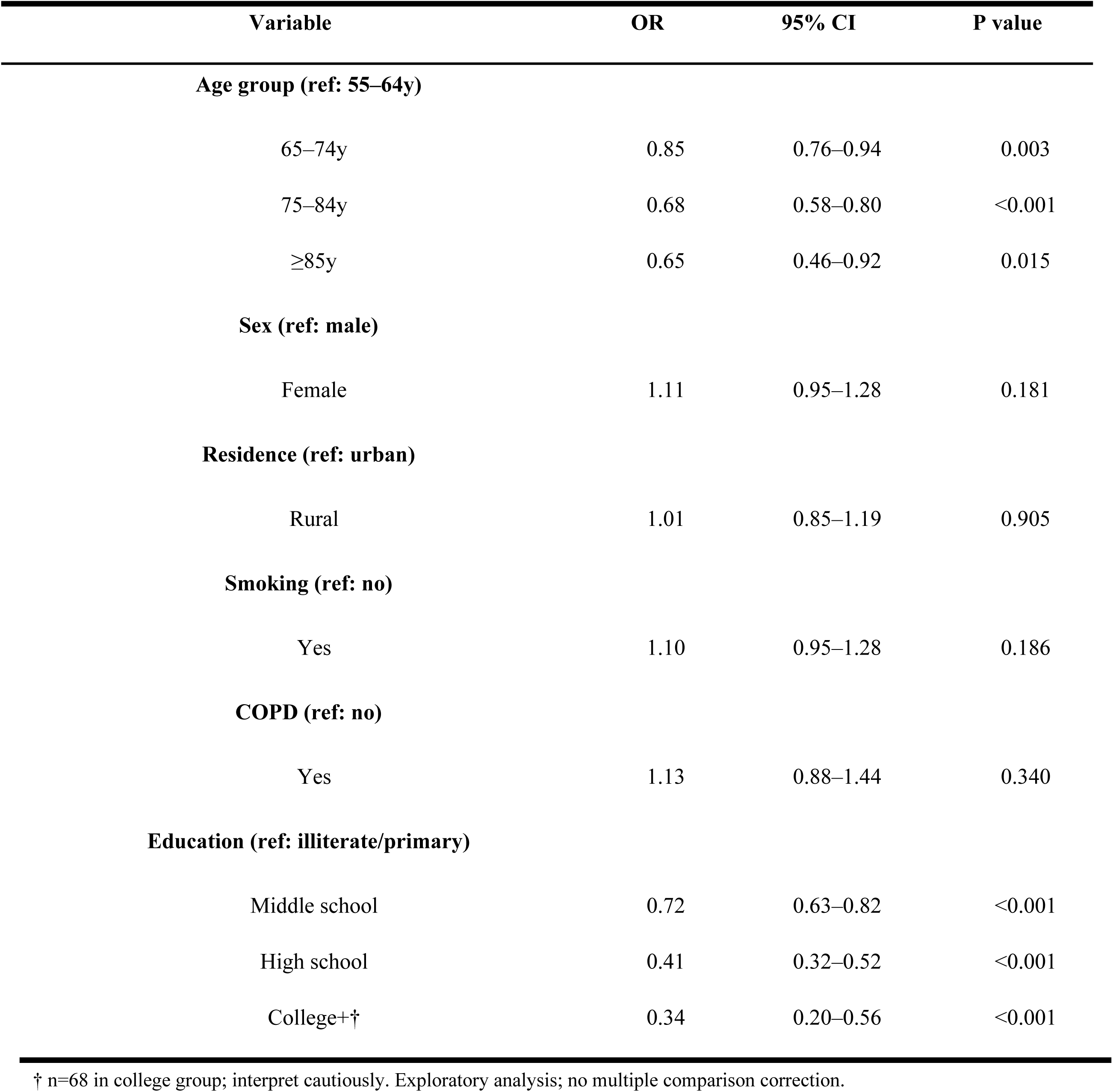
Multivariable logistic regression: factors associated with influenza vaccination (CHARLS 2020, N=13,815)

| Variable | OR | 95% CI | P value |
| --- | --- | --- | --- |
| **Age group (ref: 55–64y)** |  |  |  |
| 65–74y | 0.85 | 0.76–0.94 | 0.003 |
| 75–84y | 0.68 | 0.58–0.80 | <0.001 |
| ≥85y | 0.65 | 0.46–0.92 | 0.015 |
| **Sex (ref: male)** |  |  |  |
| Female | 1.11 | 0.95–1.28 | 0.181 |
| **Residence (ref: urban)** |  |  |  |
| Rural | 1.01 | 0.85–1.19 | 0.905 |
| **Smoking (ref: no)** |  |  |  |
| Yes | 1.10 | 0.95–1.28 | 0.186 |
| **COPD (ref: no)** |  |  |  |
| Yes | 1.13 | 0.88–1.44 | 0.340 |
| **Education (ref: illiterate/primary)** |  |  |  |
| Middle school | 0.72 | 0.63–0.82 | <0.001 |
| High school | 0.41 | 0.32–0.52 | <0.001 |
| College+† | 0.34 | 0.20–0.56 | <0.001 |

† n=68 in college group; interpret cautiously. Exploratory analysis; no multiple comparison correction.

### GBD-CHARLS Ecological Analysis

The correlation analysis between disease burden and vaccination coverage across age groups yielded a pattern that was hard to miss. Plotting GBD 2020 age-specific mortality rates against CHARLS 2020 vaccination coverage for the nine age groups gave a Spearman correlation of −0.700 (p = 0.036). In other words, the groups with the highest mortality risk were consistently those with the lowest vaccine uptake—a pattern we ended up calling a “protection mismatch.” We ran the same analysis using GBD 2023 data to see if this was just a one-year artifact; it came back at nearly the same estimate (r = −0.700, p = 0.036), so the mismatch appears to be a persistent feature rather than a temporary blip. The full correlation matrix is in Table 5, and Figure 5 visualizes the age-specific relationship.

**Figure 5.**
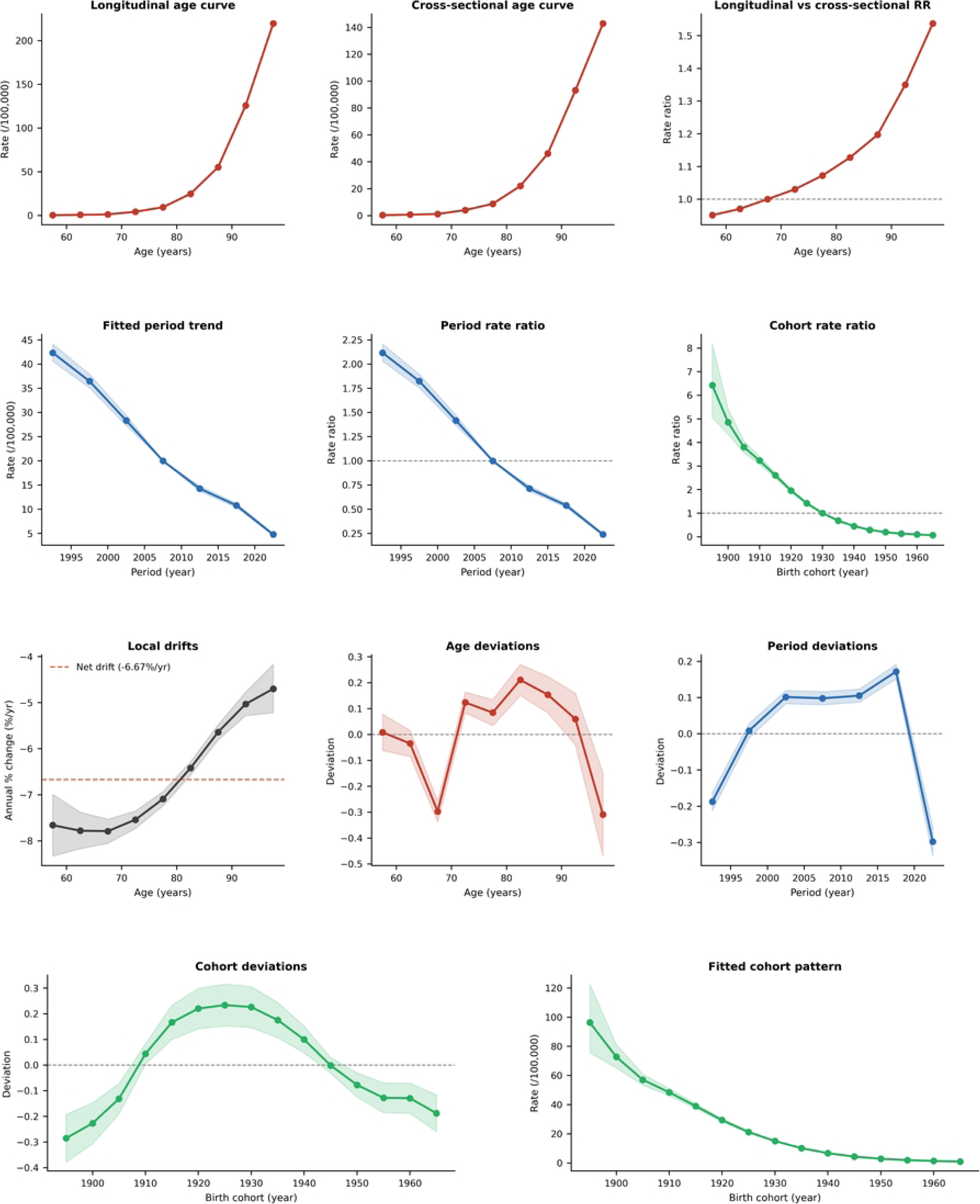
Ecological scatter plots: GBD 2023 age-specific LRTI mortality vs CHARLS 2020 indicators. A: vs vaccination coverage (r=−0.700, p=0.036); B: vs COPD prevalence.

**Table 5.** Age-specific mortality rates (GBD 2023) and CHARLS 2020 indicators by age group: ecological analysis.

| Age group | n | Mortality rate<sup>†</sup><br>(/100,000) | Vaccination (%) | COPD (%) |
| --- | --- | --- | --- | --- |
| 55–59y | 3,183 | 0.08 | 16.1 | 4.0 |
| 60–64y | 3,329 | 0.17 | 15.9 | 4.5 |
| 65–69y | 3,170 | 0.29 | 15.5 | 4.4 |
| 70–74y | 1,943 | 0.98 | 14.8 | 5.5 |
| 75–79y | 1,198 | 2.09 | 11.4 | 5.5 |
| 80–84y | 654 | 5.26 | 14.1 | 6.7 |
| 85–89y | 267 | 11.02 | 13.5 | 3.0 |
| 90–94y | 58 | 22.25 | 10.3 | 10.3 |
| ≥95y | 13 | 34.13 | 15.4 | 7.7 |

Spearman $r=-0.700$ , $p=0.036$ ( $n=9$ groups). <sup>†</sup> 2023 GBD LRTI mortality. Ecological design cannot infer individual causality.

What these ecological data point to, we think, is a particularly vulnerable subgroup. Adults aged 75 and older face what we have come to describe as a “triple vulnerability“—they carry the heaviest mortality burden, have the lowest vaccination coverage, and, as we showed earlier, also have the highest prevalence of COPD. Taken together, these converging risk factors make this age group the most obvious target for focused interventions.

### Preventable Death Estimation

We then estimated the potential mortality impact of scaling up vaccination coverage to the WHO-recommended target. Assuming coverage among adults aged ≥55 years could be raised from the current 15.1% to 75%, the number of deaths that might be averted annually ranges from 1,632 under a conservative vaccine efficacy assumption to 2,041 under a moderate scenario and 2,449 under an optimistic scenario. These figures translate to 18.7%, 23.4%, and 28.1%, respectively, of the total 5,808 deaths recorded in 2023. What stood out when we stratified these estimates by age was the overwhelming contribution of the oldest groups—adults aged 75 years and older would account for 82.7% of all preventable deaths across the three scenarios (900, 1,125, and 1,349 deaths, respectively). The detailed age-stratified estimates are provided in Table 6, and Figure 6 visualizes the projected reductions across age groups under each efficacy scenario. These findings, to our minds, make a fairly compelling case for prioritizing the oldest age groups in any future vaccination strategy.

**Figure 6.**
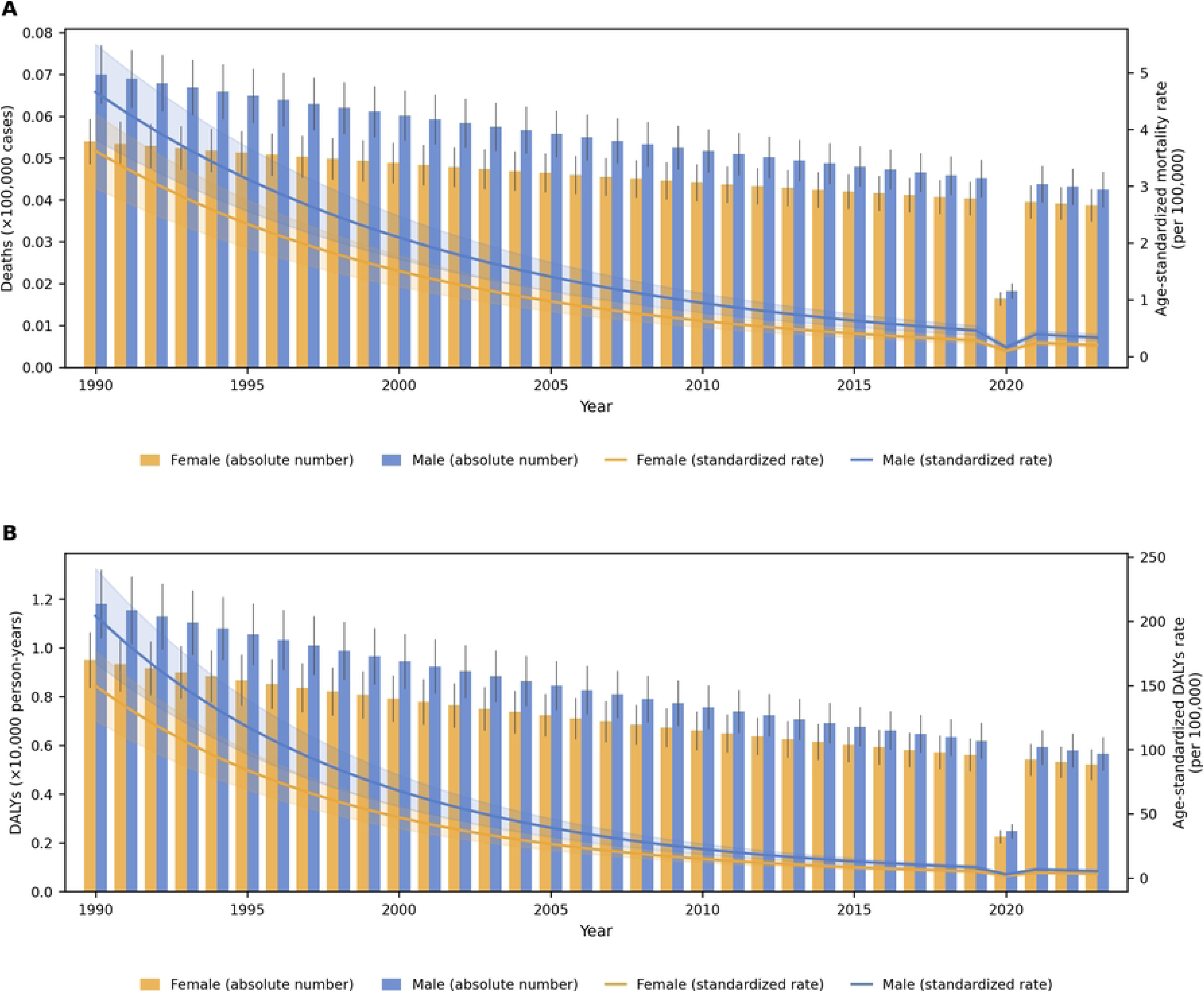
Estimated preventable influenza-associated LRTI deaths if vaccination coverage increased to 75%, by age group and VE scenario. Yellow shading: ≥75-year population.

**Table 6.**
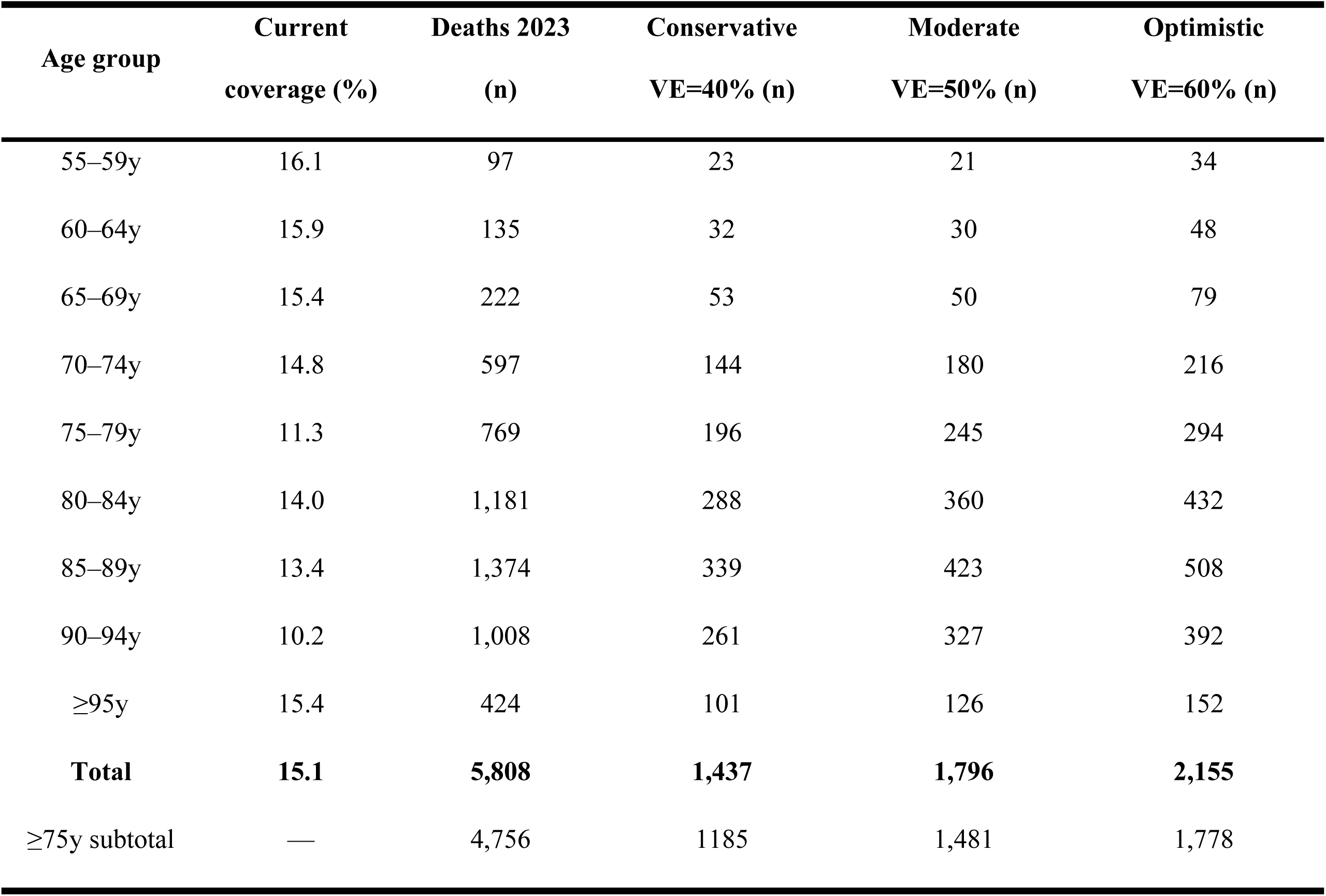
Estimated preventable deaths from influenza-associated LRTI if vaccination coverage increased to 75%, by age group (2023)

| Age group | Current coverage (%) | Deaths 2023 (n) | Conservative VE=40% (n) | Moderate VE=50% (n) | Optimistic VE=60% (n) |
| --- | --- | --- | --- | --- | --- |
| 55–59y | 16.1 | 97 | 23 | 21 | 34 |
| 60–64y | 15.9 | 135 | 32 | 30 | 48 |
| 65–69y | 15.4 | 222 | 53 | 50 | 79 |
| 70–74y | 14.8 | 597 | 144 | 180 | 216 |
| 75–79y | 11.3 | 769 | 196 | 245 | 294 |
| 80–84y | 14.0 | 1,181 | 288 | 360 | 432 |
| 85–89y | 13.4 | 1,374 | 339 | 423 | 508 |
| 90–94y | 10.2 | 1,008 | 261 | 327 | 392 |
| ≥95y | 15.4 | 424 | 101 | 126 | 152 |
| **Total** | **15.1** | **5,808** | **1,437** | **1,796** | **2,155** |
| ≥75y subtotal | — | 4,756 | 1185 | 1,481 | 1,778 |

## DISCUSSION

This study demonstrates substantial declines in influenza-associated LRTI burden among Chinese adults aged ≥55 years from 1990 to 2023, with age-standardized mortality and DALYs rates declining >96%. These improvements align with global trends but exceed worldwide averages, likely attributable to China’s rapid healthcare system development, widespread availability of neuraminidase inhibitors [23], improved nutritional status, and public health infrastructure investments [24].

A pronounced negative period deviation in 2020—a 59.6% decline in absolute deaths relative to expected trends—provides compelling ecological evidence that non-pharmaceutical interventions (NPIs) deployed during COVID-19 substantially interrupted influenza transmission, independent of vaccination. Key measures included universal masking (>95% compliance in urban areas), school and venue closures, gathering restrictions, and intensified hand hygiene campaigns.[25–27] Chinese National Influenza Center surveillance documented near-complete suppression of seasonal influenza activity during 2020–2021, with laboratory-confirmed cases declining >99% and test positivity dropping from 15–25% to <1%. Similar NPI-mediated suppression occurred globally—New Zealand (96% hospitalization reduction), Hong Kong (89%), the UK (91%), and South Korea (83%)—suggesting that targeted behavioral interventions, particularly masking and distancing, could complement vaccination during periods of high influenza activity or vaccine constraints. However, caveats remain: NPIs carry substantial economic and social costs, public compliance waned after restrictions lifted, and suppressed circulation may reduce population immunity, creating “immunity debt”. [28] Thus, while NPIs demonstrate biological efficacy, vaccination remains the sustainable, cost-effective cornerstone of prevention.

Alarmingly, CHARLS 2020 data show influenza vaccination coverage among Chinese adults aged ≥55 years at only 15.1%—over fivefold below the WHO target of ≥75% for older adults.[29,30]This coverage deficit reflects systemic failures in vaccine supply chains, financing, provider recommendation practices, and public communication, not mere individual choice. Comparator upper-middle-income countries with lower GDP per capita achieve higher coverage: Brazil ∼35% (subsidized public program integrated into primary care), Mexico ∼30% (targeted social security campaigns), and Thailand ∼25% (community health center outreach), underscoring that policy choices—not national wealth—drive vaccination gaps.

A particularly concerning finding is the inverse association between older age and vaccination uptake (OR=0.65 for ≥85 vs 55–64 years, p=0.015), exemplifying “protection-risk inversion“—those at highest risk are least likely to receive preventive care.[31]. Physical barriers include limited mobility, transport constraints, and lack of caregiver accompaniment, especially relevant in China where home-based vaccination is rare and family physician systems remain underdeveloped.[32] Among CHARLS respondents aged ≥85 years, 42% reported difficulty walking 1 km unassisted, and only 18% lived within 1 km of a vaccination-capable facility. Physician-related factors include lower recommendation rates for frail elderly due to misperceptions about blunted immunogenicity and safety concerns, despite evidence of 30–50% effectiveness in adults ≥75 years and rare serious adverse events. Notably, COPD status did not predict vaccination (OR=1.13, 95% CI: 0.88–1.44, p=0.340), indicating missed opportunities despite COPD being a clear guideline indication. Patient-related factors encompass fatalistic attitudes, perceptions of influenza as trivial, low health literacy, and vaccine safety skepticism amplified by social media misinformation[33,34].

International experiences offer solutions. The UK achieves >70% coverage in adults ≥65 years through proactive GP invitation letters, opportunistic vaccination during chronic disease visits, community pharmacy-based services, care home mandates, and sustained public campaigns. [35,36] Pharmacist-led models in the US and Australia have increased coverage by 10–15 percentage points. Adapting these to China—via community health center home visits in urban areas, village clinic campaigns integrated with annual health exams in rural areas, and systematic vaccination prompts in chronic disease management software—warrants pilot testing through stepped-wedge cluster randomized trials.

The paradoxical inverse association between higher education and vaccination (OR=0.34 for college-educated vs illiterate/primary school, 95% CI: 0.20–0.56, p<0.001) requires cautious interpretation given the very small college-educated sample (n=68, 0.5% of the analytic sample). If robust, this may represent a “health paradox“[37,38] wherein highly educated individuals prefer alternative prevention, exhibit skepticism toward pharmaceutical interventions, or rely on internet-sourced information that amplifies safety concerns. However, the sparse-data problem means this estimate carries wide uncertainty, and the association might reverse in larger samples—a question for future studies.

Ecological analysis linking GBD age-specific mortality rates with CHARLS vaccination coverage revealed a strong negative correlation (Spearman r=−0.700, p=0.036, n=9), consistent across GBD 2020 and 2023 data. Mortality risk spanned a 425-fold gradient (0.08 to 34.13 per 100,000), yet coverage remained nearly flat (15.4–16.1%), visually confirming the absence of risk-stratified vaccination targeting. While ecological analyses cannot establish individual-level causality due to ecological fallacy and potential confounding, their convergence with individual-level regression findings substantially strengthens the “protection mismatch” conclusion through triangulation[39]. Future multilevel models and natural experiments leveraging regional policy variation could improve causal inference.

Preventable death estimates indicate that raising coverage from 15.1% to the WHO-recommended 75% target would prevent approximately 1,632–2,449 deaths annually under conservative-to-optimistic vaccine efficacy assumptions. Under a moderate VE=50% scenario, the estimated 2,041 preventable deaths represent 23.4% of the 2023 influenza-associated LRTI death toll—comparable to stroke prevention via blood pressure control (∼25%) and lung cancer prevention via smoking cessation (∼30%). Notably, 82.7% of preventable deaths occur in adults ≥75 years, and the 75–79 age group has the lowest coverage (11.3%) and largest coverage gap (48.7 percentage points). Home-based vaccination, long-term care facility mandates, and integration into geriatric chronic disease management would disproportionately benefit these high-risk groups. Cost-effectiveness studies from Canada ($10,000–15,000/QALY) and the Netherlands (€12,000/QALY) support the economic viability of older adult vaccination [40],preliminary Chinese estimates suggest ∼¥8,000/QALY (≈$1,200), well below the GDP-based threshold (¥240,000/QALY).

Future research priorities include: (1) prospective cohort studies to establish temporally ordered causal relationships between barriers and vaccination behavior; (2) real-world VE studies using CHARLS-linked insurance data for China-specific estimates by age, comorbidity, and vaccine type; (3) comprehensive health technology assessments incorporating direct and indirect costs; (4) COPD-specific vaccine effectiveness studies in Chinese populations; and (5) pragmatic cluster randomized trials comparing delivery models with embedded process evaluations.

Limitations include the cross-sectional CHARLS design precluding causal inference; a three-year temporal gap between CHARLS 2020 and GBD 2023 data (though coverage and age structure remained relatively stable); an ecological analysis with only nine age groups, limiting power and vulnerable to outliers; preventable death estimates relying on literature-derived VE assumptions without accounting for annual strain matching or implementation constraints; self-reported data subject to recall bias; and multiple hypothesis testing increasing Type I error risk. These findings should be interpreted as exploratory, generating hypotheses for confirmatory studies.

## CONCLUSIONS

Influenza-associated LRTI burden in Chinese adults aged ≥55 years has fallen sharply over the past three decades — yet the oldest age groups have seen the slowest gains, and vaccination coverage remains far below what is needed. A persistent “protection mismatch” runs through the data: those at greatest risk of dying are least likely to be vaccinated. Adults aged ≥75 years sit at the intersection of high mortality, low coverage, and elevated COPD comorbidity — a triple vulnerability that makes them the logical first target for any serious effort to translate improved coverage into lives saved.

## Data Availability

CHARLS was approved by the Biomedical Ethics Committee of Peking University (IRB00001052-11015) all participants provided informed consent. This study used publicly available de-identified data requiring no additional ethical review. GBD 2023 data are publicly available with no ethical restrictions.

https://charls.pku.edu.cn/? https://vizhub.healthdata.org/gbd-results/

## ABBREVIATIONS

APC: Age-period-cohort
CHARLS: China Health and Retirement Longitudinal Study
CI: Confidence interval
COPD: Chronic obstructive pulmonary disease
DALYs: Disability-adjusted life years
EAPC: Estimated annual percentage change
GBD: Global Burden of Disease
LRTI: Lower respiratory tract infection
NPIs: Non-pharmaceutical interventions
OR: Odds ratio
UI: Uncertainty interval
VE: Vaccine efficacy
WHO: World Health Organization

## Ethics approval and consent to participate

This study utilized de-identified data from the China Health and Retirement Longitudinal Study (CHARLS), which was approved by the Biomedical Ethics Committee of Peking University (IRB00001052-11015). All CHARLS participants provided written informed consent. The Global Burden of Disease (GBD) 2023 data are publicly available and de-identified, and their use is not subject to ethical restrictions. All methods in this secondary analysis were performed in accordance with the relevant guidelines and regulations. As this study exclusively involved the analysis of publicly available, de-identified secondary data, no additional ethical approval was required.

## Consent for publication

Not applicable.

## Availability of data and materials

GBD 2023 data are publicly available at https://vizhub.healthdata.org/gbd-results. CHARLS 2020 data are available at https://charls.pku.edu.cn/ upon reasonable request following data access procedures.

## Authors’ contributions

ML: Conceptualization, Data curation, Formal analysis, Writing – original draft. RC: Supervision, Writing – review & editing. All authors read and approved the final manuscript.

## Acknowledgements

We acknowledge the Global Burden of Disease Study 2023 collaborators and the CHARLS research team for making their data publicly available.

## REFERENCES

1. Iuliano AD, Roguski KM, Chang HH, Muscatello DJ, Palekar R, Tempia S, et al. Estimates of global seasonal influenza-associated respiratory mortality: a modelling study. The Lancet. 2018 Mar;391(10127):1285–300. doi:10.1016/S0140-6736(17)33293-2

2. Hay AJ, McCauley JW. The WHO global influenza surveillance and response system (GISRS)—A future perspective. Influenza and Other Respiratory Viruses. 2018;12(5):551–7. doi:10.1111/irv.12565

3. Metersky ML, Masterton RG, Lode H, File TM, Babinchak T. Epidemiology, microbiology, and treatment considerations for bacterial pneumonia complicating influenza. International Journal of Infectious Diseases. 2012 May 1;16(5):e321–31. doi:10.1016/j.ijid.2012.01.003

4. McCullers JA. The co-pathogenesis of influenza viruses with bacteria in the lung. Nat Rev Microbiol. 2014 Apr;12(4):252–62. doi:10.1038/nrmicro3231

5. Franceschi C, Garagnani P, Parini P, Giuliani C, Santoro A. Inflammaging: a new immune–metabolic viewpoint for age-related diseases. Nat Rev Endocrinol. 2018 Oct;14(10):576–90. doi:10.1038/s41574-018-0059-4

6. Nikolich-Zugich J. The twilight of immunity: emerging concepts in aging of the immune system. Nature Immunology. 2018;19(1):10–9. doi:10.1038/s41590-017-0006-x

7. GBD 2023 Lower Respiratory Infections Collaborators. Global burden of lower respiratory infections and aetiologies, 1990-2023: a systematic analysis for the Global Burden of Disease Study 2023. Lancet Infectious Diseases. 2025.

8. National Bureau of Statistics of China. Communiqué of the Seventh National Population Census (No. 5). Beijing: National Bureau of Statistics of China; 2021.

9. Kyu HH, Vongpradith A, Sirota SB, Novotney A, Troeger CE, Doxey MC, et al. Age–sex differences in the global burden of lower respiratory infections and risk factors, 1990–2019: results from the Global Burden of Disease Study 2019. The Lancet Infectious Diseases. 2022 Nov;22(11):1626–47. doi:10.1016/S1473-3099(22)00510-2

10. Li L, Liu Y, Wu P, Peng Z, Wang X, Chen T, et al. Influenza-associated excess respiratory mortality in China, 2010–15: a population-based study. The Lancet Public Health. 2019 Sep;4(9):e473–81. doi:10.1016/S2468-2667(19)30163-X

11. Rondy M, El Omeiri N, Thompson MG, Levêque A, Moren A, Sullivan SG. Effectiveness of influenza vaccines in preventing severe influenza illness among adults: A systematic review and meta-analysis of test-negative design case-control studies. J Infect. 2017 Nov;75(5):381–94. doi:10.1016/j.jinf.2017.09.010 PubMed PMID: 28935236; PubMed Central PMCID: PMC5912669.

12. DiazGranados CA, Dunning AJ, Kimmel M, Kirby D, Treanor J, Collins A, et al. Efficacy of High-Dose versus Standard-Dose Influenza Vaccine in Older Adults. New England Journal of Medicine. 2014 Aug 14;371(7):635–45. doi:10.1056/NEJMoa1315727

13. Fan J, Cong S, Wang N, Bao H, Wang B, Feng Y, et al. Influenza vaccination rate and its association with chronic diseases in China: Results of a national cross-sectional study. Vaccine. 2020 Mar 4;38(11):2503–11. doi:10.1016/j.vaccine.2020.01.093

14. Zhao Y, Hu Y, Smith JP, Strauss J, Yang G. Cohort Profile: The China Health and Retirement Longitudinal Study (CHARLS). Int J Epidemiol. 2014 Feb 1;43(1):61–8. doi:10.1093/ije/dys203

15. Bray F, Ferlay J, Soerjomataram I, Siegel RL, Torre LA, Jemal A. Global cancer statistics 2018: GLOBOCAN estimates of incidence and mortality worldwide for 36 cancers in 185 countries. CA: A Cancer Journal for Clinicians. 2018;68(6):394–424. doi:10.3322/caac.21492

16. La Vecchia C, Levi F, Lucchini F, Negri E. Trends in mortality from major diseases in Europe, 1980–1993. Eur J Epidemiol. 1998 Jan 1;14(1):1–8. doi:10.1023/A:1007440201137

17. Yang Y, Fu WJ, Land KC. A Methodological Comparison of Age-Period-Cohort Models: The Intrinsic Estimator and Conventional Generalized Linear Models. Sociological Methodology. 2004;34(1):75–110. doi:10.1111/j.0081-1750.2004.00148.x

18. Age Period Cohort Analysis Tool [Internet]. [cited 2026 May 17]. Available from: https://analysistools.cancer.gov/apc/

19. Morgenstern H. Ecologic studies in epidemiology: concepts, principles, and methods. Annu Rev Public Health. 1995;16:61–81. doi:10.1146/annurev.pu.16.050195.000425 PubMed PMID: 7639884.

20. National Immunization Advisory Committee (NIAC) Technical Working Group (TWG), Influenza Vaccination TWG. Technical guidelines for seasonal influenza vaccination in China (2023-2024). Chinese Journal of Epidemiology. 20231012;44(10):1507–30. doi:10.3760/cma.j.cn112338-20230908-00139

21. Gross P, Hermogenes A, Sacks H. The efficacy of influenza vaccine in elderly persons: a meta-analysis and review of the literature. Annals of Internal Medicine. 1995;123(7):518–27. doi:10.7326/0003-4819-123-7-199510010-00008

22. Mulpuru S, Li L, Ye L, Hatchette T, Andrew MK, Ambrose A, et al. Effectiveness of Influenza Vaccination on Hospitalizations and Risk Factors for Severe Outcomes in Hospitalized Patients With COPD. Chest. 2019 Jan 1;155(1):69–78. doi:10.1016/j.chest.2018.10.044

23. Dobson J, Whitley RJ, Pocock S, Monto AS. Oseltamivir treatment for influenza in adults: a meta-analysis of randomised controlled trials. The Lancet. 2015 May 2;385(9979):1729–37. doi:10.1016/S0140-6736(14)62449-1 PubMed PMID: 25640810.

24. Wang L, Wang Y, Jin S, Wu Z, Chin DP, Koplan JP, et al. Emergence and control of infectious diseases in China. Lancet. 2008 Nov 1;372(9649):1598–605. doi:10.1016/S0140-6736(08)61365-3 PubMed PMID: 18930534; PubMed Central PMCID: PMC7138027.

25. Cowling BJ, Ali ST, Ng TWY. Impact assessment of non-pharmaceutical interventions against coronavirus disease 2019 and influenza in Hong Kong: an observational study. The Lancet Public Health. 2020;5(5):e279–88. doi:10.1016/S2468-2667(20)30090-6

26. Wang L, Wang Y, Jin S. Emergence and control of infectious diseases in China. Lancet. 2008;372(9649):1598–605. doi:10.1016/S0140-6736(08)61365-3

27. Huang QS, Wood T, Jelley L. Impact of the COVID-19 nonpharmaceutical interventions on influenza and other respiratory viral infections in New Zealand. Nature Communications. 2021;12(1):1001. doi:10.1038/s41467-021-21157-9

28. Cohen R, Ashman M, Taha MK. Pediatric Infectious Disease Group (GPIP) position paper on the immune debt of the COVID-19 pandemic in childhood. Infectious Diseases Now. 2021;51(5):418–23. doi:10.1016/j.idnow.2021.05.004

29. World Health Organization. Global Influenza Strategy 2019-2030 [Internet]. Geneva: World Health Organization; 2019. Available from: https://www.who.int/publications/i/item/global-influenza-strategy-2019-2030

30. Palache A, Oriol-Mathieu V, Fino M. Seasonal influenza vaccine dose distribution in 195 countries (2004-2013). Vaccine. 2015;33(47):6369–76. doi:10.1016/j.vaccine.2015.09.049

31. Lorenc T, Marshall D, Wright K. Seasonal influenza vaccination in adults: a systematic review of barriers, facilitators and interventions. BMC Health Services Research. 2020;20:86. doi:10.1186/s12913-020-4916-1

32. Yip W, Fu H, Chen AT. 10 years of health-care reform in China: progress and gaps in Universal Health Coverage. Lancet. 2019;394(10204):1192–204. doi:10.1016/S0140-6736(19)32136-1

33. Wang Q, Yue N, Zheng M. Influenza vaccination coverage of population and the factors influencing influenza vaccination in mainland China: a meta-analysis. Vaccine. 2018;36(48):7262–9. doi:10.1016/j.vaccine.2018.09.063

34. Grohskopf LA, Alyanak E, Broder KR. Prevention and Control of Seasonal Influenza with Vaccines: Recommendations of the Advisory Committee on Immunization Practices — United States, 2020-21 Influenza Season. MMWR Recommendations and Reports. 2020;69(8):1–24. doi:10.15585/mmwr.rr6908a1

35. Public Health England. Seasonal influenza vaccine uptake in GP patients: winter season 2019 to 2020 [Internet]. London: Public Health England; 2020. Available from: https://www.gov.uk/government/statistics/seasonal-flu-vaccine-uptake-in-gp-patients-winter-season-2019-to-2020

36. Isenor JE, Edwards NT, Alia TA. Impact of pharmacists as immunizers on vaccination rates: a systematic review and meta-analysis. Vaccine. 2016;34(47):5708–23. doi:10.1016/j.vaccine.2016.08.085

37. Piantadosi S, Byar DP, Green SB. The ecological fallacy. American Journal of Epidemiology. 1988;127(5):893–904. doi:10.1093/oxfordjournals.aje.a114892

38. Tu YK, Gunnell D, Gilthorpe MS. Simpson’s Paradox, Lord’s Paradox, and Suppression Effects are the same phenomenon — the reversal paradox. Emerging Themes in Epidemiology. 2008;5:2. doi:10.1186/1742-7622-5-2

39. Lawlor DA, Tilling K, Davey Smith G. Triangulation in aetiological epidemiology. International Journal of Epidemiology. 2016;45(6):1866–86. doi:10.1093/ije/dyw314

40. Feng J, Xu L, Wang J. Cost-effectiveness of seasonal influenza vaccination for pregnant women, healthcare workers, and adults 60 years and older in Beijing. Vaccine. 2019;37(13):1787–94. doi:10.1016/j.vaccine.2019.02.027

